# Pangenome-wide association study reveals selective absence of CRISPR genes (Rv2816c-19c) in drug-resistant *Mycobacterium tuberculosis*

**DOI:** 10.1101/2024.02.06.24302273

**Authors:** Nikhil Bhalla, Ranjan Kumar Nanda

**Affiliations:** Translational Health Group, International Center of Genetic Engineering and Biotechnology, New Delhi Component, Aruna Asif Ali Road, New Delhi, India.

**Keywords:** Drug resistance, GWAS, *M. tuberculosis*

## Abstract

**Background:** The drug resistance development in *Mycobacterium tuberculosis* (Mtb) is attributed to the acquisition of mutations in the drug target genes. However, the role of the differential presence of non-essential accessory genes is relatively unexplored and Pan Genome-Wide Association Study (Pan-GWAS) can identify these gene sets that could contribute to drug resistance development in Mtb.

**Methodology:** Publicly available Whole Genome Sequencing (WGS) data of clinical Mtb isolates (n=2601) from TB endemic countries (India, China, Zambia, Pakistan) were used in this study. The Mtb WGS data was de novo assembled, filtered for contamination, scaffolded into longer contigs, and functionally annotated. All analyses, including Gene repertoire and Pan-GWAS, were conducted using open-source tools, and the Benjamin Hochberg test was applied to identify genes having significant association with drug-resistant Mtb isolates.

**Results:** Out of 2601 Mtb WGS data sets, 2184 qualified as high-quality and were used for Pan-GWAS analysis (drug-resistant n=1386; drug-sensitive n=798). A set of 3784 core genes, 123 softcore genes, 224 shell genes, and 762 cloud genes were identified. Sets of 33 and 39 genes showed a positive and a negative association with drug-resistant isolates, respectively, with high significance (p-value < 0.01). Gene ontology cluster analysis indicated a compromised bacterial immune system and impaired DNA repair in drug-resistant compared to the sensitive isolates. Multidrug efflux pump repressor genes (Rv3830c and Rv3855c) were also absent in the drug-resistant Mtb isolates. The absence of CRISPR-associated genes (Rv2816c-19c) is reported in other drug-resistant microbes, and a similar pattern is observed in Mtb.

**Conclusions:** This study sheds light on Mtb genes involved in drug resistance emergence and could be helpful in better understanding of host-pathogen interactions, identification of novel drug targets and diagnostics.

## Introduction

According to the Global Tuberculosis Report 2023 by the World Health Organization (WHO), approximately 410,000 new cases of drug-resistant Tuberculosis (TB) were reported in 2022. TB has a higher prevalence in tropical regions, such as South Asia and Africa, mainly comprising poverty-stricken developing nations, compared to other parts of the world. The causative agent of TB, i.e., *Mycobacterium tuberculosis* (Mtb), is a slow-growing pathogen that mostly causes lower respiratory tract infections and evolved several intrinsic drug resistance-conferring factors including thick cell wall, lipid-rich cell membrane, drug-inactivating enzymes and drug target modification systems. Host-dependent selective pressures such as inappropriate treatment or inappropriate dosing of antibiotics and extrinsic drug resistance-conferring factors lead to mutations in the drug target, drug-activating genes, and their promoter regions, leading to ineffective drug action (1,2,3,4). Despite the introduction of several novel and repurposed drugs for the treatment of drug-resistant TB, it is alarming to note that TB cases unresponsive to new therapeutics are increasingly being reported (5). In addition to its slow growth rate, its ability to remain dormant for decades, the formation of a persister population and the development of drug tolerance are the reasons that can cause a high prevalence of relapse TB cases. Understanding the evolution of drug resistance is essential for the development of diagnostics, effective therapeutics, and is critical for a better understanding of host-pathogen interactions of Mtb.

Drug resistance in Mtb is primarily associated with the acquisition of Single Nucleotide Polymorphisms (SNPs) in the drug target genes, prodrug-activating genes, their promoter regions, and other genes that are involved in the mechanism of action of respective drugs. Unlike in other bacteria, Horizontal Gene Transfer (HGT) of the drug resistance-conferring plasmids do not contribute to the development of drug resistance in Mtb (6–8). However, Mtb is susceptible to infection by *Mycobacteriophages* and can also undergo natural intra-genome recombination events (9,10). In a recent study, the insertion sequence IS6110 that encodes a transposase is found to actively take part in transposition, thereby leading to genetic deletions in the observable time frame of one year. IS6110 is also reportedly more abundant in Lineage 2 (Beijing) Mtb isolates, which are widely known to be highly drug-resistant (11,12). Events like gene deletions can potentially contribute to the fitness of drug-resistant Mtb isolates, thereby leading to the emergence of drug resistance.

Genome-Wide Association Studies (GWAS) have been extensively used to identify drug resistance-associated SNPs in Mtb, but their application at the pangenome level remains limited (13). Existing literature on Pan-GWAS of Mtb seems to be primarily focused on identifying the genetic determinants taking part in higher prevalence, the site of infection (Extrapulmonary/Pulmonary) and those forming inter-species diversity (14–16). A Pan-GWAS study from 2018 identified 24 novel genetic signatures associated with drug resistance using a sample set of 1595 from varying geographical regions (17). Another Pan-GWAS study, also from 2018, was more focused on understanding the causation of atypical drug resistance in Mtb isolates. In this, several unique genes, as well as intergenic regions, are found to be exclusively associated with atypical drug resistance in Mtb compared to 145 other isolates with typical drug-resistant markers (18).

Given the limited literature on the pangenome of drug-resistant Mtb isolates and our understanding of genomic structural variations that may arise upon drug resistance development, we aimed to identify unique gene repertoires by analysing a publicly available Mtb WGS data set from TB endemic countries. The identified gene sets in the present study might be useful in developing region-specific diagnostic tools, identifying novel drug targets and understanding the host-pathogen interactions of drug-resistant Mtb isolates.

## Methodology

### Data acquisition, De novo assembly, and Metagenome detection

From existing reports with corresponding NCBI-BioProjects PRJNA879962, PRJEB41116, PRJEB32684, PRJNA379070, and PRJEB29435, Mtb WGS data (n=2601) from India, China, Pakistan, and Zambia were downloaded from the SRA-NCBI database using fasterq-dump v2.11.3 of the SRA Toolkit (NCBI) (19–23). Megahit assembler v1.2.9. was used for de novo assembly of the Mtb WGS data (24). The Ragtag tool v2.1.0. was used for scaffolding Megahit assemblies (25). Kraken2 tool v2.1.3. was used for the detection of metagenomes using Minikraken database (https://ccb.jhu.edu/software/kraken/dl/minikraken_20171019_8GB.tgz) (26). The de novo assembly quality was evaluated using the QUAST tool v5.2.0.(27).

### Drug resistance profiling, Gene Repertoire analysis, and Statistics

For drug resistance profiling and lineage identification of the Mtb isolates, the Megahit assemblies were used as input in the TBprofiler tool v5.0.0. (Database: n0599ccdEJody) and the output data was compiled using the "collate" argument (28,29). Guided functional annotation of de novo scaffolds was performed employing the Prokka tool v1.14.6. and Mtb H37Rv as refrence genome (Accession: GCF_000195955.2) as input for “--proteins” argument (30). The Gene Repertoire analysis was carried out by employing Panaroo v1.3.4. (31). Gene Ontology clustering was performed using DAVID (https://david.ncifcrf.gov/).

#### Statistical analysis

Benjamin-Hochberg test was used for analysing the association of genes to drug-sensitive and resistant Mtb isolates using the Scoary tool v1.6.16. (32). The genes qualifying the criteria of the adjusted p-value (< 0.01) and Log_10_Odds ratio (> 0.5 or < -0.5) were considered to have significantly perturbed association either with drug-resistance or drug-sensitive isolates. GraphPad Prism v8.0.2. and MS Excel (2016 home edition) were used for data analysis and representation.

## Results

### WGS data analysis, filtering and population structure

The clinical Mtb isolates (n=2601) used for this analysis were reported from India (n=2232), China (n=201), Pakistan (n=80), and Zambia (n=88) (S2-Table 1). Based on TBprofiler analysis, the total Mtb isolates were sub-grouped as drug-sensitive (n=863), rifampicin-resistant (n=90), isoniazid-resistant (n=147), mono/poly-resistant (n=117), MDR (n=465), preXDR (n=883), and XDR (n=36). The classification of isolates into drug-resistant categories (MDR, XDR, and pre-XDR) was done according to WHO 2020 recommendations (https://www.who.int/publications/i/item/9789240018662). In 76 Mtb isolates, we detected with more than one lineage of Mtb, indicating strain mixing, and the rest did not show signatures of strain mixing (n=2525, k1) (Figure 1-A and 1S-A). Based on Metagenome analysis, a subset of samples (n=365) had > 5% of total alignment with non-Mtb organisms (that included Non-tuberculous *Mycobacteria* viz., *M. kansasii, M. chimera, M. marinum*, etc and other *Actinobacteria, Proteobacteria, Firmicutes, Chlamydiae, Crenarcheota* species) and rest of them (n=2236, k2) had ≥ 95% alignment specifically with MTBC species in the Minikraken database (Figure 1A and S-1B).

**Figure 1:**
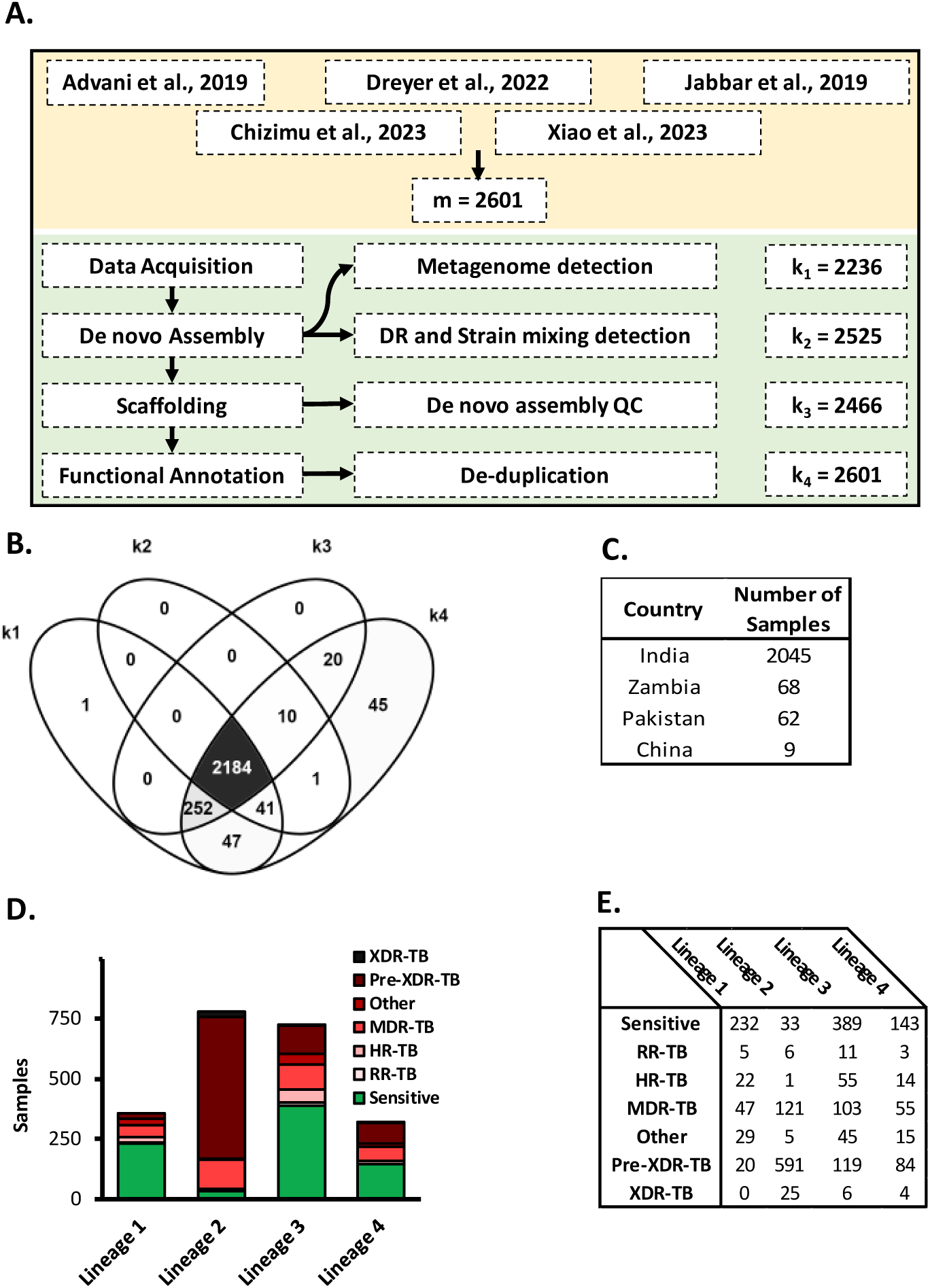
Whole Genome Sequencing (WGS) data filtering and population structure of clinical *Mycobacterial tuberculosis* isolates included in the study. **A:** Workflow employed to analyse publicly available WGS data. The data analysis consisted of data acquisition, assembling, DR profiling, lineage determination, metagenome detection, scaffolding, de novo assembly QC, functional annotation, and removal of duplicate GFF files. 2601 (m) samples were downloaded for data analysis, and k_1-4_ denotes the number of samples that successfully underwent analyses and showed high quality on various metrics. Upon Venn analysis, 2184 samples were found common in k1-4 and were subjected to gene repertoire analysis **(B)**. The sample set (2184) consisted of Mtb isolates from 4 countries **(C)** with varying drug resistance profiles and four lineages **(D and E)**. Abbreviations: DR: Drug Resistant; m: the total number of samples downloaded from NCBI-SRA; k_1_: number of samples passing >95% of reads aligning with MTBC species; k_2_: number of samples having successfully characterized with no evidence of mixed lineages; k_3_: number of samples that passed de novo quality metrics; k_4_: number of samples passing deduplication based on MD5checksum of GFF files.

**Table 1:** Significant associations of genes with resistance and sensitivity were filtered based on BH-adjusted p-value (<0.01) and Log_10_OR (>0.5 and < -0.5). The tables show genes (locus tag), their products, and the value of Log_10_OR, indicating the direction of association (Resistance and Sensitivity). The loci in red text form RD as per the RDScan database. **A**: Genes having a negative association; B: Genes having a positive association with drug resistance. Abbreviations: RD: Region of Difference; BH: Benjamin Hochberg; OR: Odds Ratio; S, Sensitive; R, Resistant.

| **A** |  |  | **B** |  |  |
| --- | --- | --- | --- | --- | --- |
| Locus Tag | Product | Log<sub>10</sub>OR | Locus Tag | Product | Log<sub>10</sub>OR |
| Rv3855 | HTH-type transcriptional repressor EthR | -2.13 | Rv3919c | 16S rRNA (guanine(527)-N(7))-methyltransferase RsmG | 2.22 |
| Rv3856c | hypothetical protein | -2.08 | Rv1844c | 6-phosphogluconate dehydrogenase | 1.79 |
| Rv2765 | hydrolase | -1.64 | **Rv1355c** | molybdopterin biosynthesis protein MoeY | 1.40 |
| Rv3830c | TetR family transcriptional regulator | -1.42 | Rv2016 | hypothetical protein | 1.39 |
| **Rv0071** | maturase | -1.41 | Rv3383c | polyprenyl synthetase IdsB | 1.32 |
| **Rv0073** | glutamine ABC transporter ATP-binding protein | -1.40 | Rv1371 | membrane protein | 1.31 |
| **Rv0072** | glutamine ABC transporter permease | -1.37 | Rv1730c | penicillin-binding protein | 1.28 |
| **Rv2816c** | CRISPR-associated endoribonuclease Cas2 | -1.30 | Rv2318 | sugar ABC transporter substrate-binding lipoprotein UspC | 1.27 |
| **Rv2818c** | CRISPR-associated protein Csm6 | -1.29 | Rv0840c | proline iminopeptidase | 1.13 |
| **Rv2819c** | CRISPR type III-associated RAMP protein Csm5 | -1.24 | Rv0968 | hypothetical protein | 1.05 |
| **Rv2817c** | CRISPR-associated endonuclease Cas1 | -1.04 | Rv1500 | glycosyltransferase | 1.02 |
| **Rv1673c** | hypothetical protein | -0.85 | Rv2231A | ribonuclease VapC16 | 1.00 |
| **Rv1672c** | integral membrane transport protein | -0.84 | Rv0648 | alpha-mannosidase | 0.98 |
| **Rv1967** | Mce family protein Mce3B | -0.80 | Rv1255c | HTH-type transcriptional regulator | 0.84 |
| Rv1038c | ESAT-6 like protein EsxJ | -0.80 | Rv0393 | hypothetical protein | 0.69 |
| Rv1583c | phage protein | -0.78 | Rv1787 | PPE family protein PPE25 | 0.65 |
| Rv1582c | phage protein | -0.77 | Rv0394c | hypothetical protein | 0.62 |
| Rv1581c | phage protein | -0.77 | Rv2653c | toxin | 0.62 |
| Rv1578c | phage protein | -0.77 | Rv2645 | hypothetical protein | 0.62 |
| Rv1573 | phage protein | -0.77 | Rv2656c | prophage protein | 0.62 |
| Rv1576c | phage capsid protein | -0.76 | Rv2658c | prophage protein | 0.62 |
| Rv1580c | phage protein | -0.76 | Rv2657c | prophage protein | 0.62 |
| Rv1579c | phage protein | -0.76 | Rv2659c | prophage integrase | 0.61 |
| Rv1575 | phage protein | -0.75 | Rv2646 | probable integrase | 0.61 |
| Rv1585c | phage protein | -0.75 | Rv2655c | prophage protein | 0.61 |
| Rv1527c | polyketide synthase | -0.69 | Rv2650c | prophage protein | 0.60 |
| Rv1760 | diacylglycerol acyltransferase | -0.67 | Rv1577c | phage prohead protease | 0.58 |
| Rv1762c | hypothetical protein | -0.65 | Rv2652c | prophage protein | 0.58 |
| Rv1758 | cutinase | -0.64 | Rv2654c | antitoxin | 0.57 |
| Rv1213 | glucose-1-phosphate adenyltransferase | -0.64 | Rv1788 | PE family protein PE18 | 0.56 |
| Rv1761c | hypothetical protein | -0.63 | **Rv3467** | hypothetical protein | 0.54 |
| Rv1765c | hypothetical protein | -0.60 | Rv3020c | ESAT-6 like protein EsxS | 0.53 |
| Rv1557 | transmembrane transport protein MmpL6 | -0.60 | **Rv1979c** | permease | 0.52 |
| Rv2319c | universal stress protein | -0.60 |  |  |  |
| Rv3737 | transmembrane protein | -0.58 |  |  |  |
| Rv1266c | serine/threonine-protein kinase PknH | -0.57 |  |  |  |
| Rv3515c | acyl-CoA synthetase | -0.56 |  |  |  |
| Rv3798 | insertion sequence element IS1557 transposase | -0.54 |  |  |  |
| Rv0963c | hypothetical protein | -0.51 |  |  |  |

Additional parameters like Mtb genome size (∼4411532 bp), GC % (∼65), and outliers having too many mismatches compared to the reference genome was used to filter the samples. Scaffolded de novo assemblies (n=2466, k3) had GC% > 62, N50 > 3999999, genome fraction relative to Mtb H37Rv > 95 %, and mismatches per 100 kbp < 100 (Figure 1S-C). One sample failed to undergo scaffolding and was excluded. MD5Checksum of annotated genomes (GFF format) revealed that 2600 annotated genomes were non-duplicates (k4) (Figure 1A). Finally, a set of 2184 samples, qualified all four data processing and filtering steps (DR profiling and strain mixing determination, Metagenome detection, De novo QC, and Md5Checksum-based de-duplication of annotated genomes) (Figure 1B).

The high-quality sample set (n=2184) consisted samples from India (n=2045), China (n=9), Pakistan (n=62) and Zambia (n=68) (Figure 1C). A majority (95.8%) of Lineage 2 Mtb isolates were found as drug-resistant, while 55% of Lineage 4, 46.6% of Lineage 3, and 34.6% of Lineage 1 samples were determined as drug-resistant. Approximately 64% (n=1386) of these isolates were drug-resistant (25 Rif-resistant, 95 isoniazid-mono-resistant, 326 MDR, 94 mono/poly drug-resistant, 814 preXDR and 35 XDR samples), and 36% (n=798) were classified as drug-sensitive (Figure 1D, 1E and S2-Table 2). Phylogenetic analysis grouped these samples into 4 lineages and separate clades (Figure 2A).

**Figure 2:**
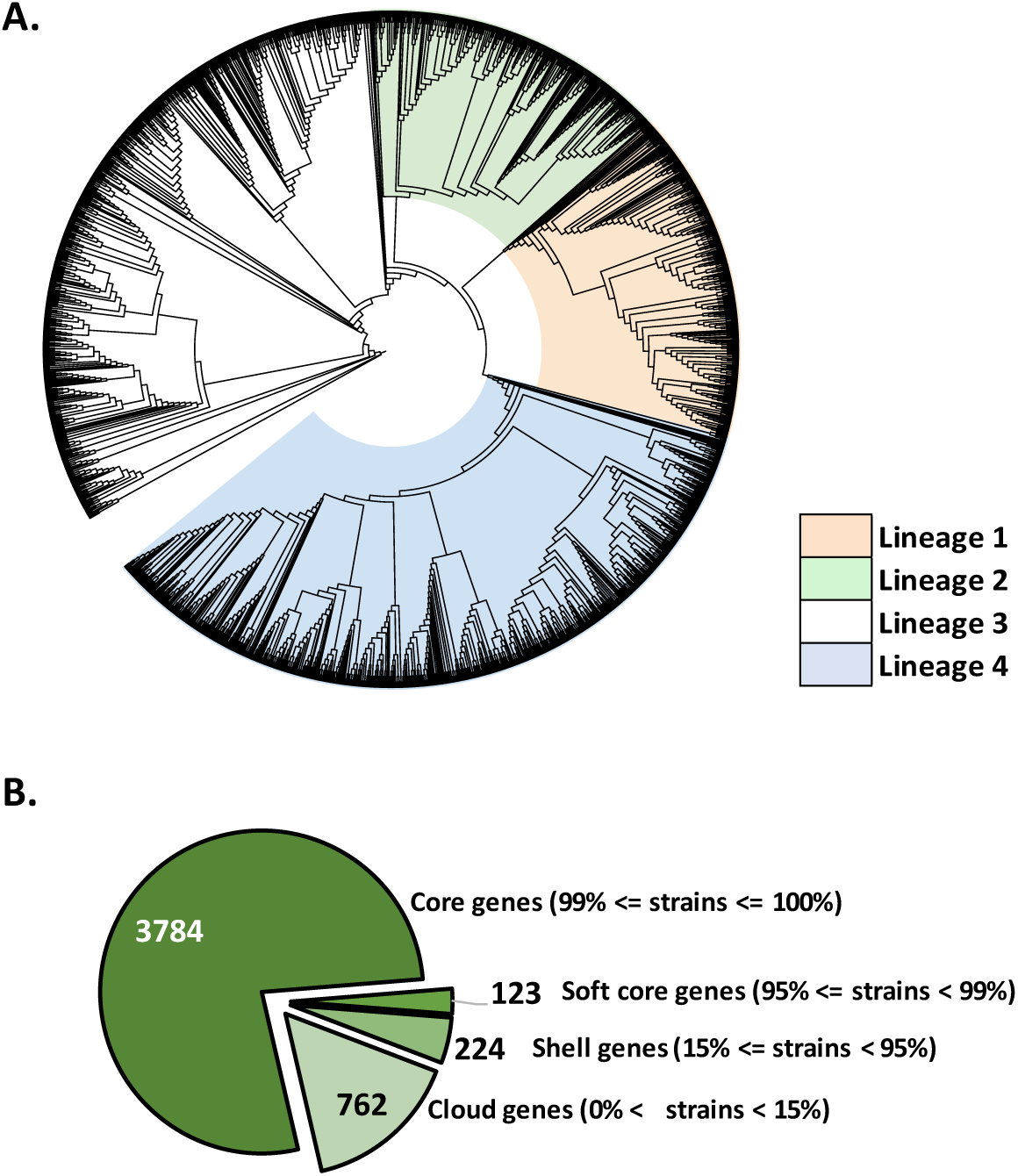
Phylogenetic analysis and summary statistics of gene repertoire (GR) analysis. **A:** Phylogenetic dendrogram of the high-quality sample set (n=2184) used for the GR analysis. The accessory gene alignment in Newick format was visualized on iTOL. The dendrogram shows four Mtb lineages as separate clades. **B:** Summary Statistics of Gene Repertoire analysis.

### Pan-GWAS analysis reveals gene repertoire differences in drug-resistant and sensitive Mtb isolates

In total, a set of 4893 genes were found in these 2184 high-quality Mtb genomes (Figure 2B). Gene repertoire analysis of the qualified sample set using Panaroo showed 3784 core genes (present in >99 % of isolates), 123 softcore genes (present in 95-98% of isolates), 224 shell genes (present in 15-94% of isolates), and 762 genes (present in >0-14% isolates)(31). A set of 187 genes was found to be significantly associated with either drug resistance or drug sensitivity status (Benjamin Hochberg adjusted p-value < 0.01). Amongst these, 115 gene sets aligned to multiple genes of Mtb H37Rv and because of their redundancy were excluded from further analysis. Out the rest 72 gene clusters, 39 genes showed a negative association with drug resistance (absent in drug-resistant isolates) while 33 genes showed a positive association (present in drug resistant isolates).

The genes having significant association with either drug resistance or sensitivity status with the RDScan database (https://github.com/dbespiatykh/RDscan/blob/master/resources/RD.bed), a set of 13 genes from the known region of differences (RD) were identified. These included Rv0071-73, Rv1355c, Rv1672c-73c, Rv1967, Rv1979c, Rv2816c-19c and Rv3467.

Eleven genetic islands, each consisting of more than one tandem gene with respect to Mtb H37Rv reference, were observed to have differential presence. Six of these islands including Rv0071-73, Rv1573-85c; Rv1672c-73c, Rv1760c-62c, Rv2816c-19c, and Rv3855-56c showed negative association with drug resistance (absent in drug-resistant isolates) and 5 genetic islands, including Rv0393-94c, Rv1787-88, Rv2318A-19c, Rv2645-46, Rv2652c-59c showed positive association with drug resistance status (i.e. absent in drug-sensitive isolates) (Table 1).

Many yet-to-be-characterized hypothetical gene clusters were observed to have specific associations with one of the groups viz., Rv0393 (Resistance), Rv0394c (R), Rv0963c (Sensitive), Rv0968 (R), Rv1761c-62c (S), Rv1765c (S), Rv2016 (R), Rv3467 (R), and Rv3856c (S). Phage protein genes (Rv1573c-85c) were observed to negatively associated with drug resistance (absent in drug-resistant Mtb isolates) and genes encoding prophage proteins Rv2655c-59c have positive association with drug resistance status (present in drug-resistant isolates). After sorting the gene clusters following the Log_10_OR in ascending order, Rv3855, Rv2765, Rv3830c, Rv0071, Rv0072 and Rv0073 were found as top 6 genes with most negative association values (of Log_10_OR). Likewise, Rv3919c, Rv1844c, Rv1355c, Rv3383c, Rv1371 were found as bottom-most 5 genes having most positive values indicating positive association with drug-resistant isolates.

Apart from these genes with extreme Log_10_OR association scores, other genes like CRISPR-associated genes (Rv2816c-19c) and Toxin-Antitoxin genes (Rv2231A: VapC16 toxin and Rv2653c-54c) showed negative and positive association with drug resistance status respectively, with high significance. The genes with significant association with either drug resistance or sensitivity are compiled in Table 1A and 1B.

### Gene Ontology analysis

DAVID gene ontology (GO) clustering of the genes present only in the drug-sensitive Mtb isolates, showed enriched biological processes like antiviral defense (GO 0051607 and KW0051), endonuclease activity (KW0255), nuclease activity (KW0540), and Hydrolases (KW0378) (Figure 3A). This indicates the bacterial immune system is lost in most drug-resistant isolates.

**Figure 3:**
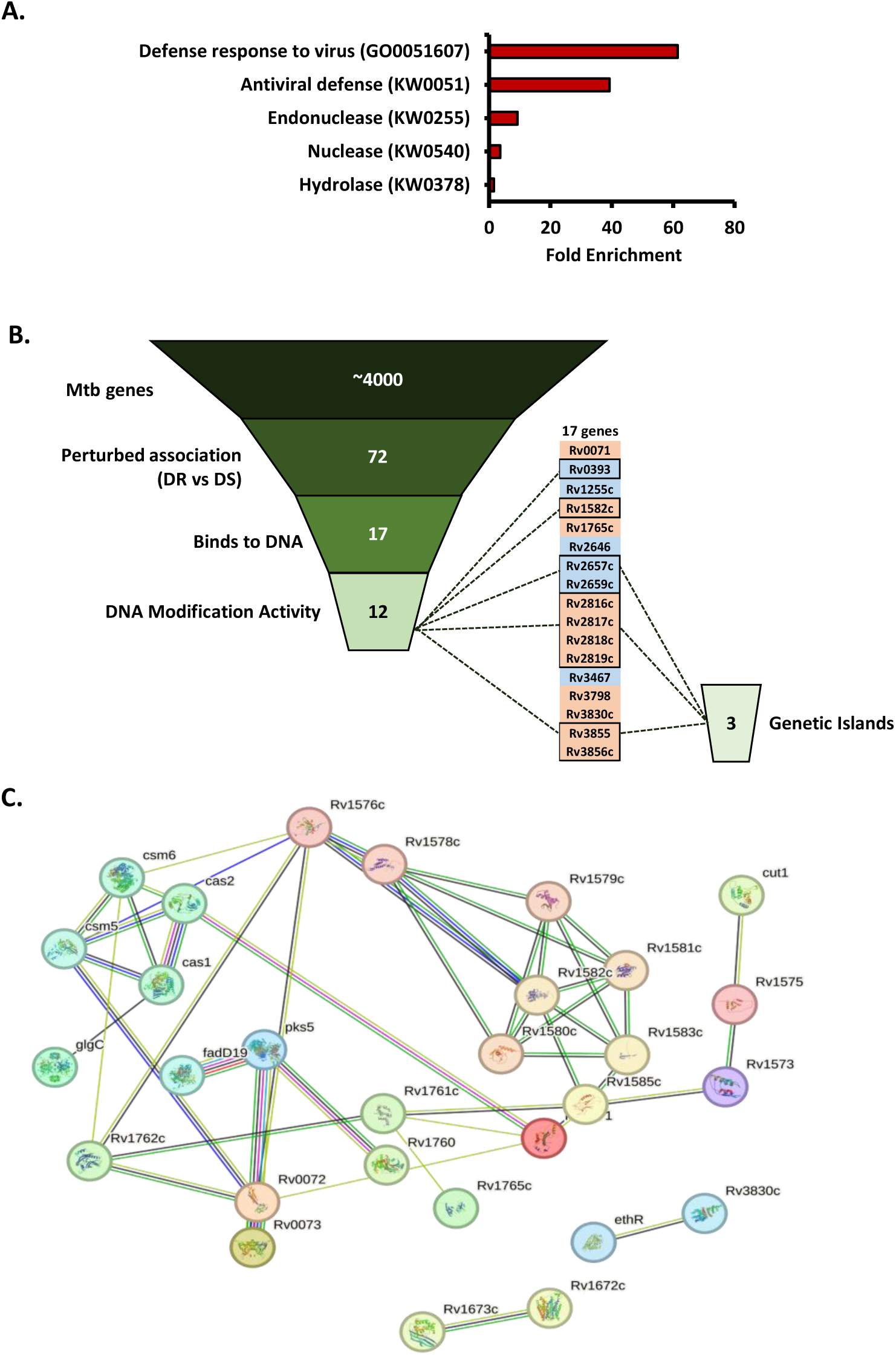
Gene ontology clustering and relationships between deregulated genes. **A**: Gene ontology clustering revealed enrichment of genes involved in inter-related GO biological processes (Defense response to invading viruses, symbionts, and other organisms). **B**: Identifying genes potentially taking part in DNA modification. Pan-GWAS revealed 72 highly significant gene associations with drug-resistant and drug-sensitive isolates. 17 out of 72 genes encode domains that bind to DNA. 12 of 17 genes with DNA binding domains can cause DNA modification. Out of 12, 3 genetic islands were identified that contained tandem genes. **C**: StringDB network analysis revealed 3 clusters of genes having variable interactions: gene neighborhood (Green), gene fusions (Red), gene co-occurrence (Blue), text-mining (Yellow), co-expressing (Black), and protein homology (Light Blue).

GO analysis also revealed 17 out of 72 genes with perturbed association encode DNA binding domains (viz., HTH and HNH) taking part in DNA modification or gene regulation. Out of 17 genes with DNA binding domains, 12 genes encode domains (Nuclease, Primase, and Helicase) that can take active part in DNA modification. Out of 12 encoding DNA-modifying genes, 10 genes from 3 genetic islands were observed. Island 1 consisted of Rv2646-59c, Island 2 consisted of Rv2816c-19c, and Island 3 consisted of Rv3855-56c (Figure 3B). Most of these (Rv0071, Rv1582c, Rv1765c, Rv2646, Rv2657c-59c, Rv2816c-19c, Rv3467, Rv3798, Rv3830c) were found to have Insertion sequences and/or repetitive DNA in close proximities (< 4 kb).

## Discussion

Understanding the emergence of drug resistance in Mtb is quintessential to predicting its future evolutionary path. Under selective pressure, Mtb acquires mutations in specific genes that contribute to the drug resistance development (3). These mutations are used as markers for diagnosing the drug resistance status of clinical Mtb isolates and are detected by molecular tools. Many databases such as TBDreamDB, WHO mutation catalogue and PhyResSE provide the details of such drug resistance associated mutations (33–35). Apart from these mutations, larger deletions have the potential to contribute to the development of drug resistance emergence in Mtb. These deletions may occur upon *Mycobacteriophage*-Mtb encounters and intragenomic recombination events (9,10,36). The Mtb genome houses a plethora of intermittently as well as continuously dispersed repetitive sequences such as insertion sequences, and direct long and short repeats that are analyzed by existing molecular typing tools such as IS6110-RFLP, Spoligotyping, MIRU-VNTR, and PGRS-RFLP (36). Some of these redundant genetic elements may be present in close proximities and are reportedly involved in genome rearrangements and deletions in specific Mtb isolates (37–39). Mtb genome also houses genes such as IS6110 (IS and Transposase), Rv3798 (probable transposase as per Mycobrowser) and RD1 region having genes with transposase-like activity (40). Factors like *Mycobacteriophages*-Mtb encounters, insertion of redundant genetic elements and Transposases make Mtb vulnerable to natural recombination events that can cause gene duplications and partial or complete deletion of genetic islands consisting of more than one non-essential gene. Many of these gene islands serve as regions of differences and aid in the identification of infecting strains in surveillance studies and confer virulence status among Mtb lineages (41). Specific genes such as those that form membrane proteins and Secretome (for example, RD155 genes including *eccCb1, PE35, esxB, esxA, eccD1, espK,* and P1cA-C) are also reported to be associated with virulence in Mtb (41,42). These factors formed the foundations of this study, which involved the investigation of genes having differential presence-absence patterns in drug-resistant Mtb isolates. We speculated that the genes with differential presence-absence patterns can potentially provide clues to understanding the evolution and emergence of drug resistance in Mtb.

In this study we conducted Pan-GWAS using the publicly available WGS data of Mtb clinical isolates reported from TB endemic countries (India, Pakistan, Zambia and China). The presence of non-Mtb metagenomes in unprocessed WGS data can introduce foreign genes and affect the accuracy of the Pan-GWAS results so samples having signatures of non-Mtb metagenomes were excluded. The WGS data which had high de-novo assembly quality and were non-duplicates were included in this study (n=2184) (Figure 1). Majority (>95%) of the lineage 2 samples showed drug-resistant profiles (Figure 1D, S1-A) corroborating earlier reports (43–45). Gene repertoire analysis idenitfied 3784 genes that form the core Mtb genome, which is within the reported range (15).

After computation of Log_10_OR values to determine the direction of association of specific genes having high significance (p-value < 0.01), we observed many intricacies that can be further investigated to understand the mechanistic aspects of drug resistance development as well as associated virulence. For example, the observed pattern in phage (negatively associated with drug resistance) and prophage proteins (positively associated with drug resistance) in this study in Mtb isolates may be responsible for the previously known complex relationship between virulence and drug resistance observed in various bacteria, including in Mtb (46). We also observed positive association of Toxin-Antitoxin genes (Rv2231A, Rv2653c and Rv2654c) with drug resistance. These Mtb genes are reported to be involved in the induction of dormancy, drug tolerance and formation of persister Mtb population (47).

The Mtb genes with identical presence-absence patterns in certain genetic islands indicates that the constituting genes are perhaps acquired simultaneously in now-extinct ancestral *Mycobacterium prototuberculosis* through historical HGT or phage infection events. These genes were subsequently found to be co-expressing (StringDB).

Upon analyzing the highly significant (p-value < 0.01) genes having extreme Log_10_OR scores, strong corroboration with existing literature was found. Rv3855, Rv2765, Rv3830c, Rv0071, Rv0072 and Rv0073 were found as top 6 genes with most negative association scores which means that they are absent in significant proportion of drug resistant isolates. Rv3855 (Log_10_OR=-2.1, HTH-type transcriptional repressor EthR), a repressor of ethionamide-activating gene Rv3854 and its absence in drug resistant isolates might not directly cause ethionamide drug resistance but could increase the tolerance to ethionamide. This tolerance could offer a survival advantage, potentially contributing to the evolution of drug resistance like phenotype (48). Mutations in the upstream region of, Rv2765 (Log_10_OR=-1.6, Hydrolase) are previously known to be associated with ethambutol drug resistance (49). Absence of Rv3830c (Log_10_OR=-1.4, TetR family transcriptional regulator), is a repressor that targets the regulatory region of various multidrug efflux pumps in drug resistant Mtb isolates can potentially cause overexpression of its targets (50). Rv0071 (Log_10_OR=-1.4, Maturase), Rv0072 (Log_10_OR=1.3, Glutamine ABC transporter permease) and Rv0073 (Log_10_OR=-1.4, Glutamine ABC transporter ATP binding protein) constitute RD105 as per RDScan database (51). The deletion of RD105 is reported earlier to be associated with drug resistance development to multiple anti-TB drugs corroborating our findings (52).

Similarly, Rv3919c, Rv1844c, Rv1355c, Rv3383c, and Rv1371 were found to have the most positive values indicating a positive association with drug-resistant Mtb isolates, which means that these genes are present in a significant number of drug-resistant isolates. Rv3919c (Log_10_OR=+2.2, 16S rRNA (guanine(527)-N(7))-methyltransferase RsmG), is previously known to be associated with low-level resistance to streptomycin (53). Rv1844c (Log_10_OR=+1.79, 6-Phosphogluconate dehydrogenase), is also previously known to be associated with Isoniazid resistance (54,55). Rv1355c (Log_10_OR=+1.4, Molybdopterin biosynthesis protein) showed positive association with drug resistance. Rv1355c and the downstream gene Rv1356c are co-expressing (as per StringDB) and Rv1356c is already known to have significantly lower expression in drug-sensitive Mtb isolates (56). Based on these factors as well as their tandem occurrence on the genome, Rv1355c and Rv1356c can be speculated to have a relationship like that of a genetic island and an operon. Rv3383c (Log_10_OR=+1.3, Polyprenyl synthetase IdsB), is known to have pyrazinamide resistance-associated mutation hotspots (57). A GWAS study previously revealed an association of Rv1371 (Log_10_OR=+1.3, a membrane protein) resistance to drugs of multiple classes, including Ethambutol, injectables, Ethionamide, Delamanid and Linezolid, corroborating with our findings (58).

Important to highlight that the seemingly critical role of CRISPR-associated genes (Log_10_OR=- 1.3 to -1.04) that showed negative association with drug resistance. Absence of Rv2816c-17c is most Lineage 2 Mtb isolates is previously known and that its absence can have a cumulative effect on the impaired DNA repair in drug-resistant Mtb isolates (59). Also, overexpression of Rv2816c is known to decrease the drug susceptibility in *M. smegmatis* and knockout strains show increased drug tolerance and drug resistance-like phenotype (59–62). Wang et al. recently discussed the association of the absent CRISPR system with drug resistance in *Klebsiella pneumoniae* (63). Similarly, its absence is also known to be associated with drug resistance in Shigella (64). Earlier reports showed that the upstream region of Rv2816c-17c-18c-19c comprises insertion sequences and direct repeats, which are vulnerable to insertion sequence-driven genome deletions (65). We found that 17 out of 72 drug-resistance associated genes encoded DNA binding motifs and 12 genes can facilitate DNA modification and repair. In close proximities to these genes, we observed many repeat sequences and transposable elements. Many genes including the hypothetical ones showed strong association with drug resistance development in Mtb and may not have a direct causation. Elucidating the role of these genes with respect to the emergence of or conferring drug resistance will potentially enrich our understanding of drug resistance evolution in Mtb.

The observed patterns of genes indicate some Mtb isolates are pre-equipped to evade antibiotic treatment.

## Data Availability

All data used in the present study was obtained from the NCBI-Sequence Repository Archive. The Bioproject accession numbers are mentioned in the manuscript.

## Contributions

NB conceptualized the study and analysed data. RKN provided guidance for the execution of analysis in the study. NB and RKN prepared the manuscript.

## Acknowledgement

NB and RKN acknowledge the Department of Biotechnology (DBT), India for Funding (Grant ID: National Network Project of National Institute of Immunology, New Delhi -[40267]). Nidhi Yadav and Ashish Gupta from TH Group, ICGEB, New Delhi component are acknowledged for providing critical feedback and reviewing the manuscript.

