## Supplementary S1 for "Pangenome-wide association study reveals selective absence of CRISPR genes (Rv2816c-19c) in drug-resistant *Mycobacterium tuberculosis*"

**Supplementary 1**

***Corresponding author(s):**

Nikhil Bhalla (Ph.D.)

**or,**

Ranjan Kumar Nanda (Ph.D.)

Translational Health Group, International Centre for Genetic Engineering and Technology, New Delhi-110067, India

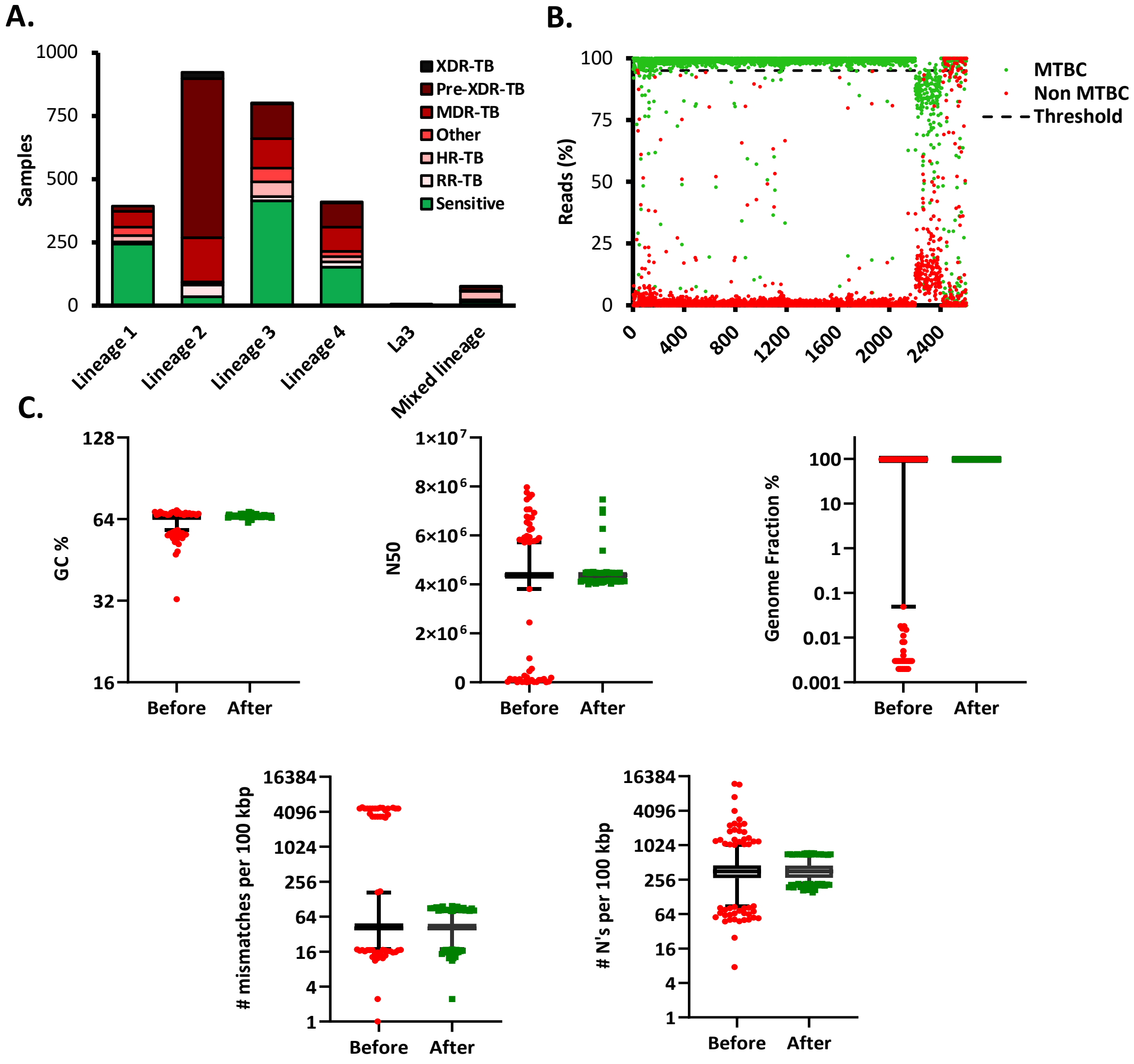


**Figure S1: QC of WGS data and filtering. A**: Percentage and Number of drug-resistant and drug-susceptible Mtb isolates. **B**: Metagenome determination in WGS data. The X-axis shows the percentage of reads aligning to MTBC (Green) and Unclassified + Non-MTBC (Red). Samples having 95% reads aligning to MTBC species were retained (Black line threshold). **C**: Quality metrics of de novo scaffolds before and after filtering (GC % > 62; N50 > 3999999; Genome Fraction % > 95; Mismatches per 100Kbp < 100).
